# Long-term mortality and cause-specific death after non-cardiac chest pain: a multicentre cohort study of 160,245 patients in China

**DOI:** 10.64898/2026.06.15.26355724

**Authors:** Yayu You, Hongchun Hu, Li Yin, Junxuan Sang, Renhe Yu, Xiuqin Hong, Yuan Liu, Fen Liu, Wen Su, Shanshan Jiang, Ying Tang, Yu Zhang, Hongwei Pan, Yan Cao, Zhengyu Liu

## Abstract

**Background:** Non-cardiac chest pain (NCCP) is commonly regarded as a low-risk condition. However, long-term mortality, cause-specific death, and high-risk subgroup characteristics remain poorly defined.

**Methods:** In this multicentre registry-linked cohort study, we linked the Chest Pain Center Registry from 101 hospitals in Hunan, China, with the Mortality and Cause of Death Registry. Adults diagnosed with NCCP from Jan 1, 2017, to Dec 31, 2021, were included. We assessed 3-year all-cause, cardiovascular, and non-cardiovascular mortality using Cox, restricted cubic spline, and Fine-Gray models.

**Findings:** Among 160,245 patients, 4674 deaths occurred within 3 years (2.9%). Mortality increased sharply after 60.5 years. Age ≥ 60.5 years (adjusted hazard ratio [aHR] 7.49 [95% CI 6.89-8.14]), rural residence (time-varying aHR 1.46 [1.35-1.57] in year 1 and 1.66 [1.46-1.89] in years 1-3), and male sex (aHR 1.47 [1.38-1.57]) independently predicted death. Three-year mortality ranged from 0.3% in younger urban women to 8.4% in older rural men. Cardiovascular diseases accounted for 56.4% of deaths among older patients, whereas other non-cardiovascular causes (22.8%) and malignancy (20.8%) were the largest categories among younger decedents.

**Interpretation:** NCCP is not uniformly benign. Age, rural residence, and sex identify patients who could benefit from risk-stratified follow-up, with cardiovascular prevention prioritised for older rural men and broader non-cardiovascular assessment considered for younger patients.

**Research in context:** *Evidence before this study:* We searched PubMed from database inception to June 4, 2026, without date restrictions or language filters, using combinations of “non-cardiac chest pain”, “noncardiac chest pain”, “non-specific chest pain”, “nonspecific chest pain”, and “undiagnosed chest pain” with “prognosis”, “mortality”, “long-term”, “cause-specific”, and “cardiovascular mortality”. We considered systematic reviews and cohort studies reporting prognosis, mortality, cardiovascular outcomes, recurrent health-care use, or cause-specific outcomes after NCCP, non-specific chest pain, or undiagnosed chest pain. Previous evidence, including cohort studies with several years of follow-up, suggests that patients with NCCP or related chest pain diagnoses are not a homogeneous low-risk group. However, definitions, clinical settings, and outcomes have varied substantially, and cause-specific mortality has not been consistently reported. In our search, we did not identify large multicentre registry-linked studies examining long-term cause-specific mortality after NCCP or how age, sex, and urban-rural context shape risk after chest pain centre discharge.

*Added value of this study:* This multicentre registry-linked cohort included 160,245 patients with NCCP from 101 chest pain centres in Hunan, China, linked to a mortality and cause-of-death registry. The study provides 3-year estimates of all-cause, cardiovascular, and non-cardiovascular mortality, characterises cause-of-death composition, and uses competing-risk methods to separate cardiovascular from non-cardiovascular death. It also shows that simple variables available at presentation, age, sex, and residence, identify a steep mortality gradient, with older rural men having the highest absolute risk.

*Implications of all the available evidence:* The available evidence and our findings support reframing NCCP as a potential entry point for risk-stratified follow-up rather than as reassurance alone. Age, sex, and residence could help chest pain services and primary care systems identify patients who need closer post-discharge surveillance. The urban-rural gradient also points to continuity of care and access to cardiovascular prevention as public health priorities after chest pain evaluation. Follow-up strategies should be aligned with competing risks: cardiovascular prevention may be most relevant for older patients, whereas broader assessment of non-cardiovascular conditions may be appropriate for younger patients. Future studies should test whether targeted follow-up pathways can improve long-term outcomes after NCCP.

## Introduction

Non-cardiac chest pain (NCCP) imposes a substantial burden on emergency departments and outpatient clinics worldwide[1]. Although traditionally regarded as a low-risk, benign condition, emerging evidence indicates that certain NCCP subgroups carry elevated long-term cardiovascular risks and mortality[2]. A systematic review documented marked heterogeneity: some cohorts showed excess cardiovascular mortality, particularly in older patients and those with cardiovascular risk factors, whereas others found no differences from the general population[3]. A critical limitation is that most studies have reported only all-cause mortality, without distinguishing cardiovascular from non-cardiovascular cause[2, 3]. It therefore remains unclear whether NCCP patients predominantly die from cardiovascular disease or from competing non-cardiovascular conditions. Moreover, the long-term prognostic profile of NCCP, including which patients are at highest risk and which clinical features define them, has not been systematically examined in large population-based studies.

This knowledge gap is especially relevant where demographic and geographic factors drive wide outcome disparities. In China, urban-rural gaps in healthcare access and chronic disease care, compounded by rapid aging, mean that the prognostic significance of NCCP likely differs substantially across patient groups[4, 5]. Using a large-scale cohort of 160,245 NCCP patients followed for a median of 3 years, we characterized the long-term prognostic landscape within a competing risks framework. Specifically, we quantified mortality risks by age, residence, and sex, described the predominant causes of death across these strata, and identified subgroups in whom NCCP could serve as a trigger for risk-stratified follow-up and stronger linkage between chest pain services and primary care.

## Methods

### Data sources

This multicentre, registry-linked, retrospective cohort study used data from two integrated provincial-level databases in China: the Chest Pain Center Registry (CPCR), a standardised registry of 101 hospitals (91 certified by the Chinese National Chest Pain Center plus 10 applicant hospitals) that prospectively collects demographics, clinical characteristics, and ICD-10-coded discharge diagnoses for all patients presenting with chest pain; and the Mortality and Cause of Death Registry (MCDR), a province-wide vital statistics system maintained by the Center for Disease Control and Prevention that records date and underlying cause of death. Deterministic linkage between the two registries was performed using unique resident identification numbers. The study was approved by the Ethics Committee of Hunan Provincial People’s Hospital [approval number: 2026-018] and registered at the Chinese Clinical Trial Registry (ChiCTR2500091922). Informed consent was waived owing to the retrospective design and use of de-identified data. Reporting follows the STROBE guidelines[6].

### Study population

Adults aged 18 years or older presenting between 1 January 2017 and 31 December 2021 with chest pain as the primary diagnosis (ICD-10 R07) were eligible[7]. NCCP was operationally defined as an index chest pain presentation without concurrent ICD-10 codes for acute cardiac aetiologies (ischaemic heart disease I20-I25, pulmonary embolism, aortic dissection, or other specified cardiovascular conditions; details in supplementary table). Patients with a documented history of cardiovascular disease or cancer (C00-C97) before the index visit were excluded. After applying exclusion criteria, 160,245 patients were included. Follow-up commenced at the index visit and continued until death or 3 years, whichever occurred first.

### Outcomes

The primary outcome was all-cause mortality. For cause-specific analyses, cardiovascular (CV) death was defined as death attributed to diseases of the circulatory system (ICD-10 I00-I99); all other deaths were classified as non-cardiovascular (non-CV) death. For descriptive analyses, deaths were further classified into eight categories based on three-character ICD-10 prefixes: ischaemic heart disease (I20-I25), ischaemic stroke (I63), intracerebral haemorrhage (I61), other CV death, respiratory diseases (J codes), genitourinary diseases (N codes), malignant neoplasms (C codes and D00-D48), and other non-CV death (details in supplementary methods S2).

### Exposures and covariates

The primary exposure was age, dichotomised at 60.5 years (<60.5 years [reference] vs ≥ 60.5 years), with the cutoff determined by the Youden index[8] from receiver operating characteristic (ROC) analysis. Residence (urban [reference] vs rural) and sex (female [reference] vs male) were examined as additional primary exposures. Covariates included occupation (employed, unemployed, student), hypertension, diabetes, lipid-lowering drug use, smoking, and body mass index (BMI). Covariates with a standardised mean difference greater than 0.10 between exposure groups were included in multivariable models (supplementary methods S3). Co-primary exposures were mutually adjusted regardless of SMD (Table S1). Collinearity was excluded (all VIF < 2.5; supplementary methods S4). BMI replaced separate height and weight terms because of strong weight-BMI correlation.

### Statistical analysis

Continuous variables were summarised as mean (SD) and categorical variables as frequency (%). Baseline characteristics were stratified by death outcome (alive, CV death, non-CV death), with group comparisons using chi-squared and t-tests.

Age was evaluated as a predictor of all-cause mortality using ROC analysis with the Youden index to determine the optimal dichotomisation threshold. The continuous age-mortality relationship was modelled using restricted cubic splines (RCS) with four knots[9], adjusted for sex, residence, occupation, hypertension, diabetes, lipid-lowering drug, smoking, and BMI, with the reference set at 60.5 years.

Kaplan-Meier survival curves were constructed by age group, age-sex strata, and age-residence strata. Multivariable Cox proportional hazards models were fitted with all three primary exposures mutually adjusted plus the full covariate set[10]. The proportional hazards assumption was tested using Schoenfeld residuals[11]. For exposures violating the assumption, follow-up was split at 1 year using survSplit to yield period-specific hazard ratios (0-1 year, 1-3 years). For cause-specific mortality, cumulative incidence functions were estimated using the Aalen-Johansen estimator, and Fine-Gray subdistribution hazard models were fitted with the same covariate set to estimate subdistribution hazard ratios (sHR) and 95% CIs[12].

The joint effects of age, residence, and sex were assessed by cross-classifying the three exposures into eight mutually exclusive subgroups, with urban female patients younger than 60.5 years serving as the reference. Cause-of-death distributions were visualised across six population subgroups among decedents. Sensitivity analyses employed incremental adjustment with six nested Cox models per exposure.

All analyses were performed using R version 4.2.3[13]. A two-sided P < 0.05 was considered statistically significant. Complete methods, including data preprocessing, ICD-10 classification, covariate selection details, collinearity diagnostics, and model specifications, are provided in the supplementary methods.

## Results

### Baseline Characteristics of the Study Population

Of the 160,245 eligible patients with non-cardiac chest pain, 155,571 (97.1%) survived, 2,503 (1.6%) died of cardiovascular (CV) causes, and 2,171 (1.4%) died of non-CV causes during 3 years of follow-up. Overall mortality was 2.9%; CV and non-CV deaths accounted for 53.6% and 46.4% of all deaths, respectively (Table 1). Compared with patients who died of non-CV causes, those who died of CV causes were older (mean age, 73.7 vs 68.6 years; P<0.001), had lower body mass index (22.7 vs 23.0 kg/m²; P=0.009), and had a higher prevalence of diabetes (10.0% vs 8.1%; P=0.026). No significant differences were observed for hypertension (52.3% vs 53.0%; P=0.644) or lipid-lowering therapy (5.0% vs 4.5%; P=0.439) (Table 1).

**Table 1:** Baseline characteristics by survival status and cause of death. Baseline characteristics of 160,245 patients with NCCP, stratified by survival status and cause of death during 3 years of follow-up. Data are mean ± SD or n (%). P values compare cardiovascular death with non-cardiovascular death, using t tests for continuous variables and chi-square tests for categorical variables.

| Characteristic |  | Overall<br>(N=160,245) | Alive<br>(n=155,571) | CV death<br>(n=2,503) | Non-CV death<br>(n=2,171) | P value<br>(CV vs Non-CV) |
| --- | --- | --- | --- | --- | --- | --- |
| Age, years | | 52.9 ± 18.2 | 52.3 ± 18.1 | 73.7 ± 11.8 | 68.6 ± 14.0 | <0.001 |
| BMI, kg/m<sup>2</sup> | | 23.3 ± 3.6 | 23.3 ± 3.6 | 22.7 ± 3.5 | 23.0 ± 3.6 | 0.009 |
| Sex | Female | 65118 (40.6%) | 63450 (40.8%) | 977 (39.0%) | 691 (31.8%) | <0.001 |
|  | Male | 95127 (59.4%) | 92121 (59.2%) | 1526 (61.0%) | 1480 (68.2%) |  |
| Residence | Rural | 73233 (45.7%) | 70491 (45.3%) | 1533 (61.2%) | 1209 (55.7%) | <0.001 |
|  | Urban | 87012 (54.3%) | 85080 (54.7%) | 970 (38.8%) | 962 (44.3%) |  |
| Career | Employed | 108310 (67.6%) | 105702 (67.9%) | 1343 (53.7%) | 1265 (58.3%) | <0.001 |
|  | Student | 8638 (5.4%) | 8631 (5.5%) | 0 (0.0%) | 7 (0.3%) |  |
|  | Unemployed | 43297 (27.0%) | 41238 (26.5%) | 1160 (46.3%) | 899 (41.4%) |  |
| Hypertension | 0 | 90917 (56.7%) | 88703 (57.0%) | 1194 (47.7%) | 1020 (47.0%) | 0.644 |
|  | 1 | 69328 (43.3%) | 66868 (43.0%) | 1309 (52.3%) | 1151 (53.0%) |  |
| Diabetes | 0 | 147865 (92.3%) | 143618 (92.3%) | 2252 (90.0%) | 1995 (91.9%) | 0.026 |
|  | 1 | 12380 (7.7%) | 11953 (7.7%) | 251 (10.0%) | 176 (8.1%) |  |
| Lipid-lowering drug | 0 | 153187 (95.6%) | 148735 (95.6%) | 2378 (95.0%) | 2074 (95.5%) | 0.439 |
|  | 1 | 7058 (4.4%) | 6836 (4.4%) | 125 (5.0%) | 97 (4.5%) |  |
| Smoking | 0 | 114918 (71.7%) | 111323 (71.6%) | 1987 (79.4%) | 1608 (74.1%) | <0.001 |
|  | 1 | 45327 (28.3%) | 44248 (28.4%) | 516 (20.6%) | 563 (25.9%) |  |

Abbreviations: BMI=body mass index; CV=cardiovascular; NCCP=non-cardiac chest pain; non-CV=non-cardiovascular; SD=standard deviation.

### Age, Residence, and Sex Were Associated With All-Cause Mortality

Receiver operating characteristic curve analysis identified 60.5 years as a data-driven threshold for risk stratification in this cohort (Figure S1A). Restricted cubic spline analysis showed a non-linear association between age and all-cause mortality (P for overall association <0.001; P for non-linearity <0.001), with risk increasing more rapidly around and beyond this threshold (Figure S1B).

In the multivariable Cox model, age ≥60.5 years, rural residence, and male sex were independently associated with higher all-cause mortality after mutual adjustment and adjustment for hypertension, diabetes, body mass index, lipid-lowering therapy, smoking, and occupation (Table 2; univariate associations in Table S2; stepwise adjustment in Table S3). Age ≥60.5 years had the strongest association (adjusted hazard ratio [aHR] 7.49; 95% CI, 6.89 to 8.14), followed by rural residence and male sex (Table 2).

**Table 2:** Associations of age, residence, and sex with all-cause mortality. Univariate and multivariable Cox proportional hazards models for all-cause mortality according to age group, residence, and sex. The multivariable model included age group, residence, and sex as co-primary exposures and was adjusted for occupation, hypertension, diabetes, lipid-lowering drug use, smoking, and body mass index. For residence, which violated the proportional hazards assumption, follow-up was split at 1 year to estimate period-specific hazard ratios.

| Exposure | Univariate HR (95% CI) | Multivariable HR (95% CI) | 0-1y HR (95% CI) | 1-3y HR (95% CI) |
| --- | --- | --- | --- | --- |
| Age ≥ 60.5y<sup>†</sup> | 8.46 (7.84-9.13) | 7.49 (6.89-8.14) | 7.73 (7.03-8.50)<sup>†</sup> | 6.77 (5.78-7.93)<sup>†</sup> |
| Rural | 1.68 (1.59-1.78) | 1.50 (1.40-1.61) | 1.46 (1.35-1.57) | 1.66 (1.46-1.89) |
| Male<sup>†</sup> | 1.23 (1.16-1.31) | 1.47 (1.38-1.57) | 1.52 (1.42-1.64)<sup>†</sup> | 1.30 (1.15-1.48)<sup>†</sup> |

<sup>†</sup> Schoenfeld test $P > 0.05$ ; proportional hazards assumption not violated. Time-varying hazard ratios shown for completeness.
Abbreviations: CI=confidence interval; HR=hazard ratio.

Schoenfeld residual testing indicated violation of the proportional hazards assumption for residence (χ²=33.75; P<0.001) (Table S4), a time-varying covariate was applied. The aHR for rural residence was 1.46 (95% CI, 1.35 to 1.57) during the first year and 1.66 (95% CI, 1.46 to 1.89) during 1-3 years of follow-up. Male sex was also independently associated with higher all-cause mortality (aHR 1.47; 95% CI, 1.38 to 1.57) (Table 2). Kaplan-Meier curves showed clear survival separation by age-sex and age-residence strata, with lower survival among older patients and additional separation by sex and residence within age groups (Figure 1). These three variables were then cross-classified for joint risk profiling.

**Figure 1:**
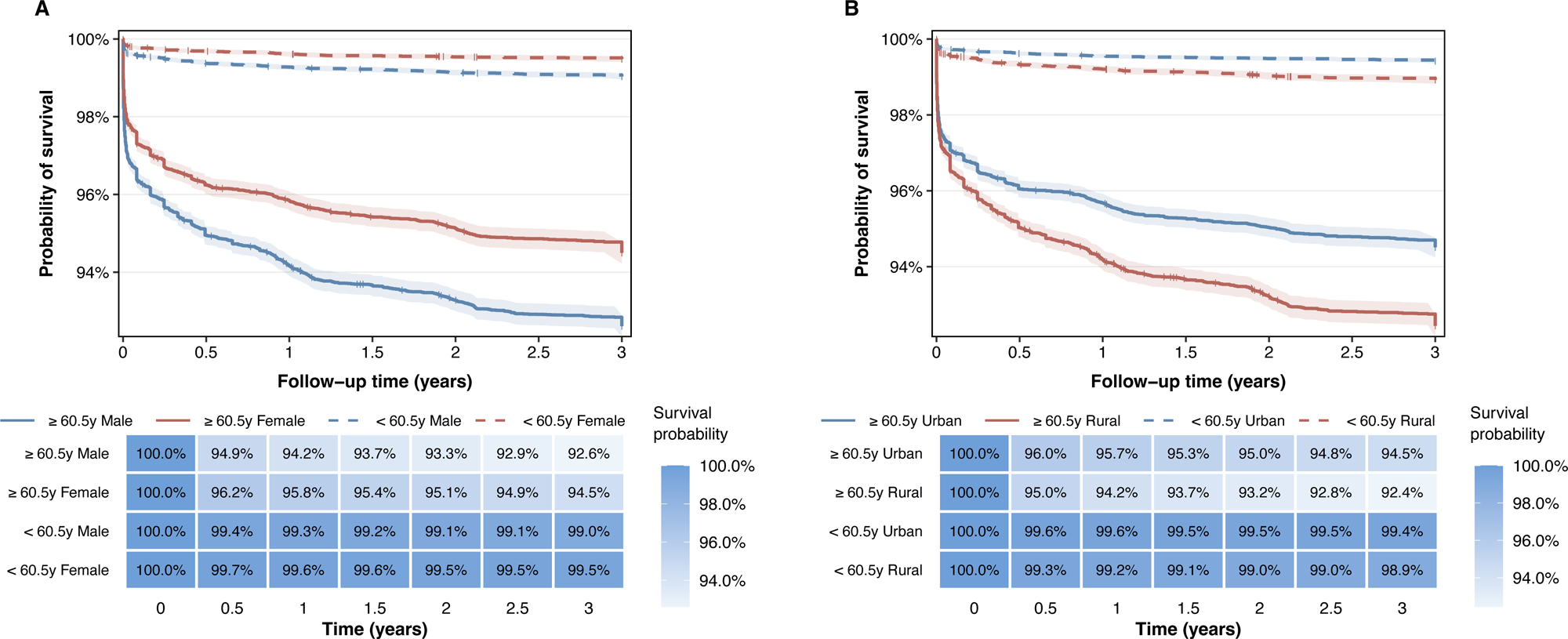
All-cause survival by age, sex, and residence. Kaplan-Meier curves for 3-year all-cause survival among 160,245 patients with NCCP, stratified by age and sex (A) and by age and residence (B). Shaded areas indicate 95% CIs; heatmaps show survival probabilities. Abbreviations: CI=confidence interval; NCCP=non-cardiac chest pain.

### Joint Effects of Age, Residence, and Sex Identified the Highest-Risk Subgroup

Cross-classification of age, residence, and sex yielded eight strata with a clear gradient in all-cause mortality (Figure 3; Table 3). Three-year mortality ranged from 0.3% in urban women younger than 60.5 years to 8.4% in rural men aged 60.5 years or older. Intermediate groups showed stepwise increases, including 0.7% in urban men younger than 60.5 years, 1.3% in rural men younger than 60.5 years, 4.6% in urban women aged 60.5 years or older, 6.3% in urban men aged 60.5 years or older, and 6.5% in rural women aged 60.5 years or older (Table 3).

**Table 3:** All-cause and cause-specific mortality risk across joint demographic strata. One-year and 3-year mortality risks and model estimates across eight strata defined by age group, residence, and sex. Urban women younger than 60.5 years were the reference group. Hazard ratios for all-cause mortality were estimated using Cox models. Subdistribution hazard ratios for cardiovascular and non-cardiovascular death were estimated using Fine-Gray models, with the alternative cause of death treated as the competing event.

| Variable | N | 1-Year Mortality | 3-Year Mortality | HR (95% CI) | P value |
| --- | --- | --- | --- | --- | --- |
| **All-cause death** |  |  |  |  |  |
| Urban, Female, < 60.5y | 22,296 | 0.3% | 0.3% | 1.00 (Reference) |  |
| Rural, Female, < 60.5y | 15,974 | 0.6% | 0.8% | 2.25 (1.69-3.00) | <0.001 |
| Urban, Male, < 60.5y | 35,445 | 0.6% | 0.7% | 2.38 (1.84-3.09) | <0.001 |
| Rural, Male, < 60.5y | 27,272 | 0.9% | 1.3% | 4.00 (3.11-5.15) | <0.001 |
| Urban, Female, ≥ 60.5y | 13,688 | 3.5% | 4.6% | 12.42 (9.73-15.86) | <0.001 |
| Urban, Male, ≥ 60.5y | 15,556 | 5.1% | 6.3% | 17.98 (14.13-22.86) | <0.001 |
| Rural, Female, ≥ 60.5y | 13,112 | 4.9% | 6.5% | 18.40 (14.52-23.31) | <0.001 |
| Rural, Male, ≥ 60.5y | 16,902 | 6.6% | 8.4% | 25.02 (19.81-31.60) | <0.001 |
| **CV death** |  |  |  |  |  |
| Urban, Female, < 60.5y | 22,296 | 0.1% | 0.1% | 1.00 (Reference) |  |
| Urban, Male, < 60.5y | 35,445 | 0.2% | 0.3% | 2.89 (1.84-4.53) | <0.001 |
| Rural, Female, < 60.5y | 15,974 | 0.2% | 0.4% | 3.23 (2.00-5.21) | <0.001 |
| Rural, Male, < 60.5y | 27,272 | 0.4% | 0.5% | 5.56 (3.60-8.60) | <0.001 |
| Urban, Female, ≥ 60.5y | 13,688 | 2.0% | 2.6% | 20.54 (13.44-31.40) | <0.001 |
| Urban, Male, ≥ 60.5y | 15,556 | 2.5% | 3.2% | 26.16 (17.14-39.93) | <0.001 |
| Rural, Female, ≥ 60.5y | 13,112 | 3.1% | 4.2% | 36.39 (24.14-54.85) | <0.001 |
| Rural, Male, ≥ 60.5y | 16,902 | 3.5% | 4.7% | 43.13 (28.66-64.91) | <0.001 |
| **Non-CV death** |  |  |  |  |  |
| Urban, Female, < 60.5y | 22,296 | 0.2% | 0.2% | 1.00 (Reference) |  |
| Rural, Female, < 60.5y | 15,974 | 0.3% | 0.4% | 1.77 (1.23-2.55) | 0.002 |
| Urban, Male, < 60.5y | 35,445 | 0.3% | 0.5% | 2.09 (1.52-2.87) | <0.001 |
| Rural, Male, < 60.5y | 27,272 | 0.5% | 0.7% | 3.18 (2.33-4.35) | <0.001 |
| Urban, Female, ≥ 60.5y | 13,688 | 1.5% | 2.0% | 8.41 (6.11-11.56) | <0.001 |
| Rural, Female, ≥ 60.5y | 13,112 | 1.7% | 2.3% | 9.46 (7.01-12.77) | <0.001 |
| Urban, Male, ≥ 60.5y | 15,556 | 2.6% | 3.2% | 13.94 (10.24-18.96) | <0.001 |
| Rural, Male, ≥ 60.5y | 16,902 | 3.1% | 3.7% | 15.72 (11.77-20.99) | <0.001 |

Abbreviations: CI=confidence interval; CV=cardiovascular; HR=hazard ratio; non-CV=non-cardiovascular; sHR=subdistribution hazard ratio.

The combined age-residence-sex stratification identified rural men aged 60.5 years or older as the subgroup with the highest 3-year all-cause mortality. We therefore used these age-residence-sex strata to examine whether cardiovascular and non-cardiovascular mortality contributed differently across risk groups.

### Cause-Specific Mortality Differed by Age Group

Competing-risk cumulative incidence analyses showed different CV and non-CV mortality patterns by age. Among patients ≥60.5 years, the 3-year cumulative incidence of CV death exceeded that of non-CV death (3.68% vs 2.85%); among patients <60.5 years, non-CV mortality exceeded CV mortality (0.48% vs 0.32%) (Figure S3, Figure 2). This pattern was consistent across sex and residence subgroups: CV mortality predominated among older patients in all eight strata, whereas non-CV mortality was comparable to or higher than CV mortality in most younger strata. Among older rural men, 3-year cumulative incidence was 4.7% for CV death and 3.7% for non-CV death; among younger men, the corresponding incidences were 0.39% and 0.58% (Table 3, Figure 2).

**Figure 2:**
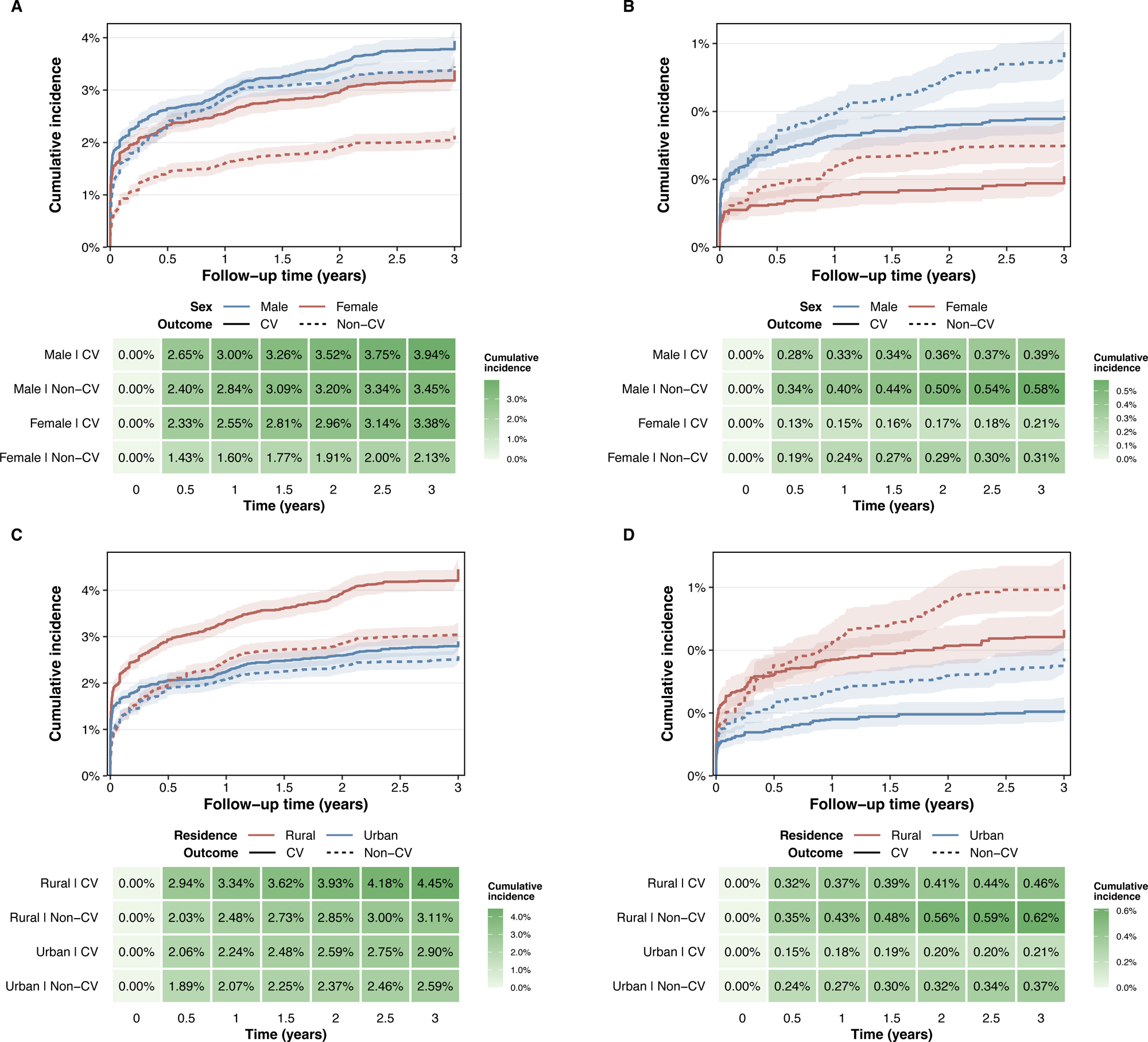
Cardiovascular and non-cardiovascular cumulative incidence by age, sex, and residence. Cumulative incidence of cardiovascular and non-cardiovascular death over 3 years by sex among older patients (A), by sex among younger patients (B), by residence among older patients (C), and by residence among younger patients (D). Older and younger patients were defined using the 60.5-year cutoff. Shaded areas indicate 95% CIs; heatmaps show cumulative incidence estimates. Abbreviations: CI=confidence interval; CV=cardiovascular; non-CV=non-cardiovascular.

Compared with urban women <60.5 years, who had a 3-year CV cumulative incidence of 0.1%, rural men ≥60.5 years had the highest risk of CV death (subdistribution hazard ratio [sHR] 43.13; 95% CI, 28.66 to 64.91), corresponding to an absolute 3-year CV cumulative incidence of 4.7% (Table 3; Figure 3). The same subgroup also had high non-CV mortality, but the relative difference from the reference group was smaller than for CV death. Full Fine-Gray sHRs for CV and non-CV death across the eight strata are reported in Table 3 and Figure 3.

**Figure 3:**
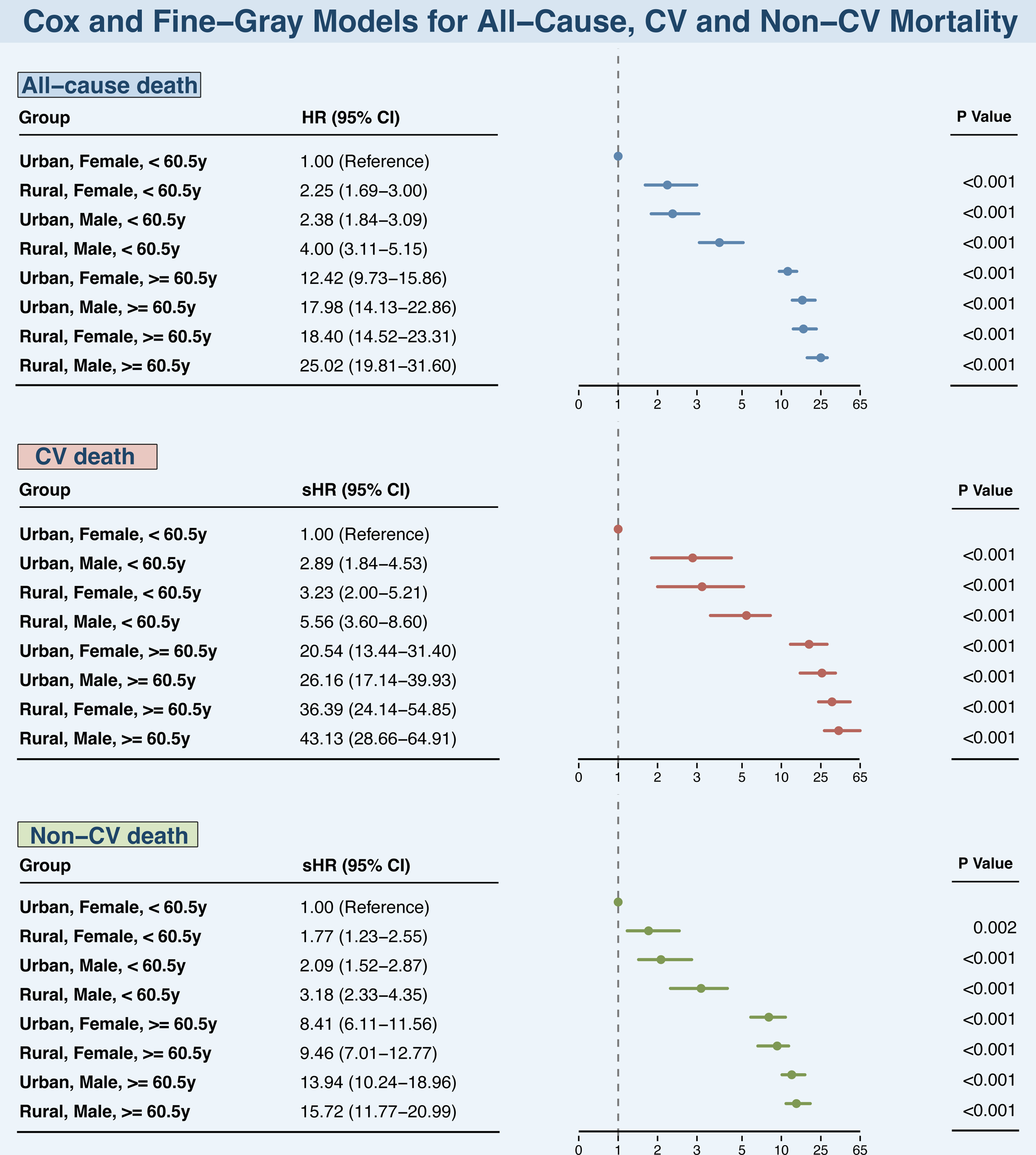
All-cause, cardiovascular, and non-cardiovascular mortality risk across joint demographic strata. Hazard ratios for all-cause death and subdistribution hazard ratios for cardiovascular and non-cardiovascular death across eight strata defined by age, sex, and residence. Urban women younger than 60.5 years were the reference group. Models were adjusted for occupation, hypertension, diabetes, lipid-lowering drug use, smoking, and body mass index. Alternative causes of death were treated as competing events in Fine-Gray models. Abbreviations: CI=confidence interval; CV=cardiovascular; non-CV=non-cardiovascular; HR=hazard ratio; sHR=subdistribution hazard ratio.

### Cause-of-Death Composition Showed Distinct Age, Residence, and Sex Patterns

Specific causes of death further differentiated mortality patterns (Figure S4, Table S5). Among decedents aged ≥60.5 years, cardiovascular diseases accounted for 56.4% of deaths; other CV causes (27.8%) and ischaemic heart disease (16.6%) were the leading specific categories. Among decedents <60.5 years, non-CV causes were more frequent; other non-CV causes (22.8%) and malignancy (20.8%) were the largest categories.

By residence, rural decedents had a higher proportion of ischaemic heart disease deaths than urban decedents (18.3% vs 13.1%) and a lower proportion of respiratory deaths (9.0% vs 15.9%). By sex, male decedents had higher proportions of malignancy (15.6%) and respiratory deaths (12.7%) than female decedents, whereas ischaemic heart disease accounted for a larger proportion of deaths in women than in men (19.8% vs 14.1%).

Overall, older rural men had the highest 3-year cumulative all-cause and CV mortality. Across age strata, CV causes predominated among older patients, whereas non-CV causes were more prominent among younger decedents, with other non-CV causes and malignancy the largest categories. Residence and sex further modified proportional cause-of-death patterns, with rural decedents showing more ischaemic heart disease and male decedents showing more malignancy and respiratory deaths. These patterns were consistent with the cumulative-incidence and cause-composition analyses shown in main Table 3, Figure 2, Figure S4, and Table S5.

## Discussion

In this large-scale cohort of 160,245 patients with non-cardiac chest pain (NCCP), prognosis over 3 years was heterogeneous rather than uniformly benign. The absolute risk gradient ranged from 0.3% in younger urban women to 8.4% in older rural men, and mortality risk increased non-linearly after approximately 60.5 years. Cause of death also differed by age: cardiovascular causes predominated among patients aged ≥60.5 years, whereas non-cardiovascular causes accounted for a larger share of deaths among patients aged <60.5 years, with other non-CV causes and malignancy the largest categories. These findings position NCCP as a potential entry point for post-discharge risk stratification. This framing is especially relevant where emergency assessment, chronic disease management, and primary-care follow-up are not consistently linked.

Previous studies have reported variable prognosis after NCCP, from low annual mortality in selected cohorts[14] to higher mortality in contemporary emergency-care populations[2]. Comparisons with acute coronary syndrome, stable coronary disease, and vascular registries provide clinical context[15–17], but long comparisons with these cohorts risk obscuring the public health question raised by NCCP. Smaller NCCP and undiagnosed chest pain studies have also signalled residual cardiac risk[18]. Our registry-linked cohort extends this evidence by combining long-term mortality, cause-specific death, competing-risk analysis, and simple demographic and geographic stratification in a large chest pain centre population.

Age was the clearest stratifying variable. The 60.5-year threshold was derived from ROC and spline analyses and is best interpreted as a pragmatic, data-driven threshold for this cohort, not as a broadly applicable clinical threshold. Its public health value lies in operational simplicity: age is available at presentation and can be used by chest pain centres and primary care teams to identify patients who might need closer cardiovascular prevention, medication review, and follow-up. The finding is consistent with the increasing absolute burden of cardiovascular and cerebrovascular death in ageing populations[19], but external validation is needed before applying this threshold in other health systems or populations.

Mortality among patients aged <60.5 years remained low and comparable to general population rates[20], whereas mortality was markedly higher among older adults, men, and rural patients. Cardiovascular deaths accounted for approximately half of deaths in older patients, consistent with accumulated cardiovascular risk and possible under-recognition of ischaemic disease after an NCCP diagnosis[21]. Among younger decedents, the larger share of non-cardiovascular deaths suggests that NCCP should not be treated as diagnostic closure when symptoms or risk factors point to non-cardiovascular disease. For older patients, residual cardiac mechanisms and later cardiovascular risk after unattributed chest pain remain relevant considerations. Rural residence should be interpreted less as an intrinsic biological risk factor than as a marker of health-system context; evidence on rural risk-factor patterns and urban-rural care differences provides context for the observed gradient[22–26].

Sex added another layer to risk stratification. Men had higher all-cause mortality, and the association strengthened after multivariable adjustment, suggesting that baseline age structure partly masked excess risk in unadjusted analyses. Men also had higher proportional mortality from malignancy and respiratory diseases[27], supporting broader non-cardiovascular assessment when clinically indicated. At the same time, ischaemic heart disease accounted for a larger share of deaths among women than among men (19.8% vs 14.1%), arguing against deprioritising cardiovascular follow-up in women after NCCP[28]. These patterns support sex-aware, rather than sex-stereotyped, post-discharge management[29], with attention to both absolute risk and cause-of-death composition.

The practical implication is a shift from a single acute exclusion pathway to a linked care pathway after chest pain evaluation. Existing protocols remain essential for ruling out acute coronary syndrome, but residual risk persists, particularly in older patients[30]. Chest pain centres could use age, residence, and sex to flag patients for risk-stratified discharge advice, communication with primary care, cardiovascular risk-factor optimisation, and timely diagnostic reassessment. For rural patients, the priority is not simply individual risk labelling but strengthening continuity of care after discharge, including referral pathways, chronic disease follow-up, and access to secondary prevention. For younger patients, follow-up pathways should also allow reassessment for non-cardiovascular disease when symptoms persist or evolve. In practical terms, this could include structured discharge summaries, clear responsibility for follow-up ownership, and prompts for cardiovascular risk review in older patients. These elements are implementation targets and their effect on outcomes would need prospective evaluation.

The main strengths of this study are its large multicentre registry-linked cohort, linkage to a mortality and cause-of-death registry, cause-specific outcome assessment, and use of competing-risk methods. The cohort design also allowed absolute risks, subgroup gradients, and cause-of-death composition to be examined within one analytic framework. These features allowed long-term risk to be described across demographic and geographic strata rather than as an average prognosis for all NCCP patients.

Several limitations warrant consideration. First, enrolment was restricted to patients presenting to certified chest pain centres, which may limit generalisability to lower-tier or non-certified settings. Second, hospital location was used as a proxy for patient residence and could have misclassified patients who crossed administrative boundaries for care, biasing urban-rural estimates toward or away from the null. Third, NCCP is a diagnosis of exclusion, and limited phenotyping for non-cardiac aetiologies may have introduced diagnostic overlap. Fourth, the absence of post-discharge data on medication use, adherence, outpatient visits, referral completion, and follow-up intensity limits assessment of care pathways after discharge. Fifth, cause-of-death coding may be imperfect, particularly for deaths occurring outside hospital settings. Finally, 3-year follow-up does not capture lifetime risk, and external validation is needed for the 60.5-year risk-stratification threshold.

In conclusion, NCCP is a heterogeneous clinical endpoint with measurable long-term mortality differences across age, sex, and urban-rural residence. Simple variables available at chest pain presentation could support risk-stratified follow-up and linkage with primary care, especially for older patients and rural populations. Future work should evaluate whether such pathways are feasible in routine chest pain services, how they should be integrated with primary care, and whether they can improve cardiovascular prevention, timely diagnostic reassessment, and long-term outcomes. In health-system terms, NCCP could become a trigger for coordinated risk review across emergency, cardiology, and primary-care services, while remaining grounded in patient symptoms, clinical judgement, and local care capacity and available resources.

## Contributors

Yayu You, Hongchun Hu, and Li Yin contributed equally to this work. Zhengyu Liu and Hongwei Pan conceived and designed the study. Yayu You, Hongchun Hu, Li Yin, and Junxuan Sang were responsible for data acquisition, data cleaning, and quality control. Yayu You and Hongchun Hu conducted the statistical analysis. Yayu You, Hongchun Hu, Junxuan Sang drafted the manuscript. Renhe Yu, Xiuqin Hong, Yuan Liu, Fen Liu, Wen Su, Shanshan Jiang, and Ying Tang contributed to data interpretation and critically revised the manuscript. Yu Zhang and Yan Cao supervised the data collection at participating hospitals. Zhengyu Liu and Hongwei Pan provided critical intellectual input and revised the final manuscript. All authors read and approved the final manuscript.

## Data Availability

Individual participant data are not publicly available because they contain sensitive health information from provincial clinical and mortality registries. Aggregated data, analysis code, and the data dictionary can be made available from the corresponding author on reasonable request after publication, subject to approval by the relevant registry data custodians and institutional review boards. Data will be shared only for scientifically sound proposals and under a signed data access agreement.

## Acknowledgments

The authors sincerely thank all participating hospitals in the Chest Pain Center Registry (CPCR) for their contributions to data collection. We are also grateful to the Disease Control and Prevention Center for providing mortality data through the Mortality and Cause of Death Registry (MCDR).

## Ethical approval

This study was registered in the Chinese Clinical Trial Registry (ChiCTR2500091922). The study protocol was approved by the Ethics Committee of Hunan Provincial People’s Hospital (The First Affiliated Hospital of Hunan Normal University) (Approval No. 2026-018). Informed consent was waived because this was a retrospective analysis of de-identified registry data.

## Data sharing

Individual participant data are not publicly available because they contain sensitive health information from provincial clinical and mortality registries. Aggregated data, analysis code, and the data dictionary can be made available from the corresponding author on reasonable request after publication, subject to approval by the relevant registry data custodians and institutional review boards. Data will be shared only for scientifically sound proposals and under a signed data access agreement. The study protocol and statistical analysis details are available in the supplementary methods.

