## Supplementary material for "Long-term mortality and cause-specific death after non-cardiac chest pain: a multicentre cohort study of 160,245 patients in China": figure and table legend ver4.2.docx

**Figure and Table Legends**

**Figure 1: All-cause survival by age, sex, and residence**

Kaplan-Meier curves for 3-year all-cause survival among 160,245 patients with NCCP, stratified by age and sex (A) and by age and residence (B). Shaded areas indicate 95% CIs; heatmaps show survival probabilities.

Abbreviations: CI=confidence interval; NCCP=non-cardiac chest pain.

**Figure 2: Cardiovascular and non-cardiovascular cumulative incidence by age, sex, and residence**

Cumulative incidence of cardiovascular and non-cardiovascular death over 3 years by sex among older patients (A), by sex among younger patients (B), by residence among older patients (C), and by residence among younger patients (D). Older and younger patients were defined using the 60.5-year cutoff. Shaded areas indicate 95% CIs; heatmaps show cumulative incidence estimates.

Abbreviations: CI=confidence interval; CV=cardiovascular; non-CV=non-cardiovascular.

**Figure 3: All-cause, cardiovascular, and non-cardiovascular mortality risk across joint demographic strata**

Hazard ratios for all-cause death and subdistribution hazard ratios for cardiovascular and non-cardiovascular death across eight strata defined by age, sex, and residence. Urban women younger than 60.5 years were the reference group. Models were adjusted for occupation, hypertension, diabetes, lipid-lowering drug use, smoking, and body mass index. Alternative causes of death were treated as competing events in Fine-Gray models.

Abbreviations: CI=confidence interval; CV=cardiovascular; non-CV=non-cardiovascular; HR=hazard ratio; sHR=subdistribution hazard ratio.

**Figure S1: Age threshold and nonlinear mortality risk**

ROC analysis identified 60.5 years as the optimal age cutoff for 3-year all-cause mortality (A). Adjusted restricted cubic spline analysis showed a nonlinear association between age and all-cause mortality, with 60.5 years as the reference (B). Shaded areas indicate 95% CIs.

Abbreviations: CI=confidence interval; ROC=receiver operating characteristic; RCS=restricted cubic spline.

**Figure S2: All-cause survival by age group**

Kaplan-Meier curves for 3-year all-cause survival, stratified by age group using the 60.5-year cutoff. Shaded areas indicate 95% CIs; the heatmap shows survival probabilities.

Abbreviations: CI=confidence interval.

**Figure S3: Cardiovascular and non-cardiovascular cumulative incidence by age group**

Cumulative incidence of cardiovascular and non-cardiovascular death over 3 years, stratified by age group using the 60.5-year cutoff. Shaded areas indicate 95% CIs; the heatmap shows cumulative incidence estimates.

Abbreviations: CI=confidence interval; CV=cardiovascular; non-CV=non-cardiovascular.

**Figure S4: Cause-of-death distribution by demographic subgroup**

Proportional distribution of underlying causes of death among decedents, stratified by age group, residence, and sex. Causes were classified using ICD-10 codes.

Abbreviations: CV=cardiovascular; ICD-10=International Classification of Diseases, 10th Revision; ICH=intracerebral haemorrhage; non-CV=non-cardiovascular.

**Table 1: Baseline characteristics by survival status and cause of death**

Baseline characteristics of 160,245 patients with NCCP, stratified by survival status and cause of death during 3 years of follow-up. Data are mean ± SD or n (%). P values compare cardiovascular death with non-cardiovascular death, using t tests for continuous variables and chi-square tests for categorical variables.

Abbreviations: BMI=body mass index; CV=cardiovascular; NCCP=non-cardiac chest pain; non-CV=non-cardiovascular; SD=standard deviation.

**Table 2: Associations of age, residence, and sex with all-cause mortality**

Univariate and multivariable Cox proportional hazards models for all-cause mortality according to age group, residence, and sex. The multivariable model included age group, residence, and sex as co-primary exposures and was adjusted for occupation, hypertension, diabetes, lipid-lowering drug use, smoking, and body mass index. For residence, which violated the proportional hazards assumption, follow-up was split at 1 year to estimate period-specific hazard ratios.

Abbreviations: CI=confidence interval; HR=hazard ratio.

**Table 3: All-cause and cause-specific mortality risk across joint demographic strata**

One-year and 3-year mortality risks and model estimates across eight strata defined by age group, residence, and sex. Urban women younger than 60.5 years were the reference group. Hazard ratios for all-cause mortality were estimated using Cox models. Subdistribution hazard ratios for cardiovascular and non-cardiovascular death were estimated using Fine-Gray models, with the alternative cause of death treated as the competing event.

Abbreviations: CI=confidence interval; CV=cardiovascular; HR=hazard ratio; non-CV=non-cardiovascular; sHR=subdistribution hazard ratio.

**Table S1: Covariate imbalance across primary exposure groups**

Standardised mean differences for candidate covariates across groups defined by residence, age group, and sex. Covariates with a standardised mean difference greater than 0.10 were included in multivariable models. Co-primary exposures were mutually adjusted regardless of standardised mean difference.

Abbreviations: BMI=body mass index; SMD=standardised mean difference.

**Table S2: Univariate predictors of all-cause mortality**

Univariate Cox proportional hazards models for all-cause mortality according to demographic characteristics, comorbidities, treatment variables, smoking, and body mass index. Age was analysed both as a binary variable dichotomised at 60.5 years and as a continuous variable per 1-year increase.

Abbreviations: BMI=body mass index; CI=confidence interval; HR=hazard ratio.

**Table S3: Incremental adjustment models for primary exposures**

Sequential Cox proportional hazards models assessing the associations of residence, age group, and sex with all-cause mortality. Models were incrementally adjusted for co-primary demographic exposures, occupation, hypertension, diabetes, lipid-lowering drug use, smoking, and body mass index.

Abbreviations: BMI=body mass index; CI=confidence interval; HR=hazard ratio.

**Table S4: Proportional hazards assumption testing**

Schoenfeld residual tests for the proportional hazards assumption in the Cox model for all-cause mortality. A significant P value indicates evidence against the proportional hazards assumption. For primary exposures violating the assumption, period-specific hazard ratios were estimated by splitting follow-up at 1 year.

Abbreviations: PH=proportional hazards.

**Table S5: Cause-of-death distribution by demographic subgroup**

Distribution of underlying causes of death among decedents during 3 years of follow-up, stratified by age group, residence, and sex. Causes of death were classified using ICD-10 codes into ischaemic heart disease, ischaemic stroke, intracerebral haemorrhage, other cardiovascular death, respiratory diseases, genitourinary diseases, cancer, and other non-cardiovascular death. Data are n (%), with percentages calculated among decedents within each subgroup.

Abbreviations: CV=cardiovascular; ICD-10=International Classification of Diseases, 10th Revision; ICH=intracerebral haemorrhage; non-CV=non-cardiovascular.
