## Supplementary material for "Long-term mortality and cause-specific death after non-cardiac chest pain: a multicentre cohort study of 160,245 patients in China": Table S1.docx

**Table S1. Covariate imbalance across primary exposure groups.**

Standardised mean differences for candidate covariates across groups defined by residence, age group, and sex. Covariates with a standardised mean difference greater than 0.10 were included in multivariable models. Co-primary exposures were mutually adjusted regardless of standardised mean difference.

| Primary exposure | Candidate covariate | SMD | Included (SMD > 0.10) |
| --- | --- | --- | --- |
| Residence | Age | 0.153 | Yes |
|  | Sex | 0.034 | Yes** |
|  | Career | 0.565 | Yes |
|  | Hypertension | 0.096 | Yes*** |
|  | Diabetes | 0.021 | No |
|  | Lipid-lowering drug | 0.093 | No |
|  | Smoking | 0.027 | No |
|  | BMI | 0.038 | No |
| Age (>= 60.5y) | Residence | 0.158 | Yes |
|  | Sex | 0.150 | Yes |
|  | Career | 0.844 | Yes |
|  | Hypertension | 0.412 | Yes |
|  | Diabetes | 0.155 | Yes |
|  | Lipid-lowering drug | 0.230 | Yes |
|  | Smoking | 0.331 | Yes |
|  | BMI | 0.197 | Yes |
| Sex | Age | 0.178 | Yes |
|  | Residence | 0.034 | Yes** |
|  | Career | 0.242 | Yes |
|  | Hypertension | 0.044 | No |
|  | Diabetes | 0.016 | No |
|  | Lipid-lowering drug | 0.040 | No |
|  | Smoking | 1.061 | Yes |
|  | BMI | 0.139 | Yes |
| SMD > 0.10 indicates covariate imbalance between exposure groups. Co-primary exposure, included for mutual adjustment regardless of SMD. Included on clinical grounds (borderline SMD, established cardiovascular risk factor). | | | |

Abbreviations: BMI=body mass index; SMD=standardised mean difference.
