## Supplementary material for "Long-term mortality and cause-specific death after non-cardiac chest pain: a multicentre cohort study of 160,245 patients in China": Table S2.docx

**Table S2. Univariate predictors of all-cause mortality.**

Univariate Cox proportional hazards models for all-cause mortality according to demographic characteristics, comorbidities, treatment variables, smoking, and body mass index. Age was analysed both as a binary variable dichotomised at 60.5 years and as a continuous variable per 1-year increase.

| Variable | HR | 95% CI | P value |
| --- | --- | --- | --- |
| Age (≥60.5y vs < 60.5y) | 8.36 | (7.62–9.16) | <0.001 |
| Age (per 1 year) | 1.07 | (1.07–1.08) | <0.001 |
| Sex (Male vs Female) | 1.23 | (1.16–1.31) | <0.001 |
| Residence (Rural vs Urban) | 1.69 | (1.58–1.80) | <0.001 |
| Student (vs Employed) | 0.03 | (0.01–0.08) | <0.001 |
| Unemployed (vs Employed) | 2.02 | (1.90–2.14) | <0.001 |
| Hypertension (Yes vs No) | 1.48 | (1.39–1.57) | <0.001 |
| Diabetes (Yes vs No) | 1.20 | (1.08–1.32) | <0.001 |
| Lipid-lowering drug (Yes vs No) | 1.09 | (0.94–1.26) | 0.252 |
| Smoking (Yes vs No) | 0.76 | (0.70–0.81) | <0.001 |
| BMI (per 1 unit) | 0.97 | (0.96–0.97) | <0.001 |

Abbreviations: BMI=body mass index; CI=confidence interval; HR=hazard ratio
