## Supplementary material for "Long-term mortality and cause-specific death after non-cardiac chest pain: a multicentre cohort study of 160,245 patients in China": Table S3.docx

**Table S3. Incremental adjustment models for primary exposures.**

Sequential Cox proportional hazards models assessing the associations of residence, age group, and sex with all-cause mortality. Models were incrementally adjusted for co-primary demographic exposures, occupation, hypertension, diabetes, lipid-lowering drug use, smoking, and body mass index.

| Exposure | Adjustment model | HR (95% CI) | P value |
| --- | --- | --- | --- |
| Residence (Rural vs Urban) | Unadjusted | 1.69 (1.58-1.80) | <0.001 |
|  | + Age + Sex | 1.45 (1.36-1.54) | <0.001 |
|  | + Career | 1.49 (1.39-1.61) | <0.001 |
|  | + HTN + DM | 1.49 (1.39-1.61) | <0.001 |
|  | + Lipid + Smoking | 1.50 (1.39-1.62) | <0.001 |
|  | + BMI (Full) | 1.50 (1.39-1.62) | <0.001 |
| Age (>=60.5y vs <60.5y) | Unadjusted | 8.36 (7.64-9.14) | <0.001 |
|  | + Age + Sex | 8.57 (7.84-9.37) | <0.001 |
|  | + Career | 8.43 (7.68-9.25) | <0.001 |
|  | + HTN + DM | 8.47 (7.69-9.31) | <0.001 |
|  | + Lipid + Smoking | 8.42 (7.64-9.28) | <0.001 |
|  | + BMI (Full) | 8.37 (7.60-9.22) | <0.001 |
| Sex (Male vs Female) | Unadjusted | 1.23 (1.16-1.31) | <0.001 |
|  | + Age + Sex | 1.42 (1.34-1.51) | <0.001 |
|  | + Career | 1.42 (1.33-1.50) | <0.001 |
|  | + HTN + DM | 1.42 (1.33-1.50) | <0.001 |
|  | + Lipid + Smoking | 1.47 (1.38-1.57) | <0.001 |
|  | + BMI (Full) | 1.47 (1.38-1.57) | <0.001 |

Abbreviations: BMI=body mass index; CI=confidence interval; HR=hazard ratio.
