## Supplementary material for "Long-term mortality and cause-specific death after non-cardiac chest pain: a multicentre cohort study of 160,245 patients in China": Table S4.docx

**Table S4. Proportional hazards assumption testing.**

Schoenfeld residual tests for the proportional hazards assumption in the Cox model for all-cause mortality. A significant P value indicates evidence against the proportional hazards assumption. For primary exposures violating the assumption, period-specific hazard ratios were estimated by splitting follow-up at 1 year.

| Variable | Chi-sq | df | P value | PH violated? |
| --- | --- | --- | --- | --- |
| Age_Risk | 2.5927122 | 1 | 0.107356356901028 | No |
| Residence | 33.7511711 | 1 | 0.000000006263133 | Yes * |
| Sex | 0.4075336 | 1 | 0.523224013989213 | No |
| Career | 17.6005420 | 2 | 0.000150692228252 | Yes * |
| Hypertension | 2.6440219 | 1 | 0.103940113304950 | No |
| Diabetes | 4.2214790 | 1 | 0.039915354821794 | Yes * |
| Lipid_lowering_drug | 4.4821987 | 1 | 0.034249632227898 | Yes * |
| smoking | 2.4495181 | 1 | 0.117560954482086 | No |
| BMI | 0.1466238 | 1 | 0.701782914729511 | No |
| GLOBAL | 55.6251306 | 10 | 0.000000024129605 | Yes * |

Abbreviations: PH=proportional hazards.
