## Supplementary material for "Long-term mortality and cause-specific death after non-cardiac chest pain: a multicentre cohort study of 160,245 patients in China": Table S5.docx

**Table S5. Cause-of-death distribution by demographic subgroup.**

Distribution of underlying causes of death among decedents during 3 years of follow-up, stratified by age group, residence, and sex. Causes of death were classified using ICD-10 codes into ischaemic heart disease, ischaemic stroke, intracerebral haemorrhage, other cardiovascular death, respiratory diseases, genitourinary diseases, cancer, and other non-cardiovascular death. Data are n (%), with percentages calculated among decedents within each subgroup.

| Subgroup | Ischaemic HD | Ischaemic stroke | ICH | Other CV | Respiratory | Genitourinary | Cancer | Other non-CV |
| --- | --- | --- | --- | --- | --- | --- | --- | --- |
| ≥60.5y (n=3,870) | 644 (16.6%) | 251 (6.5%) | 211 (5.5%) | 1075 (27.8%) | 476 (12.3%) | 145 (3.7%) | 506 (13.1%) | 562 (14.5%) |
| <60.5y (n=804) | 111 (13.8%) | 18 (2.2%) | 47 (5.8%) | 146 (18.2%) | 80 (10.0%) | 52 (6.5%) | 167 (20.8%) | 183 (22.8%) |
| Urban (n=1,942) | 254 (13.1%) | 108 (5.6%) | 97 (5.0%) | 509 (26.2%) | 309 (15.9%) | 82 (4.2%) | 258 (13.3%) | 325 (16.7%) |
| Rural (n=2,732) | 501 (18.3%) | 161 (5.9%) | 161 (5.9%) | 712 (26.1%) | 247 (9.0%) | 115 (4.2%) | 415 (15.2%) | 420 (15.4%) |
| Female (n=1,673) | 332 (19.8%) | 88 (5.3%) | 95 (5.7%) | 469 (28.0%) | 176 (10.5%) | 85 (5.1%) | 204 (12.2%) | 224 (13.4%) |
| Male (n=3,001) | 423 (14.1%) | 181 (6.0%) | 163 (5.4%) | 752 (25.1%) | 380 (12.7%) | 112 (3.7%) | 469 (15.6%) | 521 (17.4%) |

Abbreviations: CV=cardiovascular; ICD-10=International Classification of Diseases, 10th Revision; ICH=intracerebral haemorrhage; non-CV=non-cardiovascular.
