## Supplementary figures and images for "Long-term mortality and cause-specific death after non-cardiac chest pain: a multicentre cohort study of 160,245 patients in China"

### Figure S1.pdf

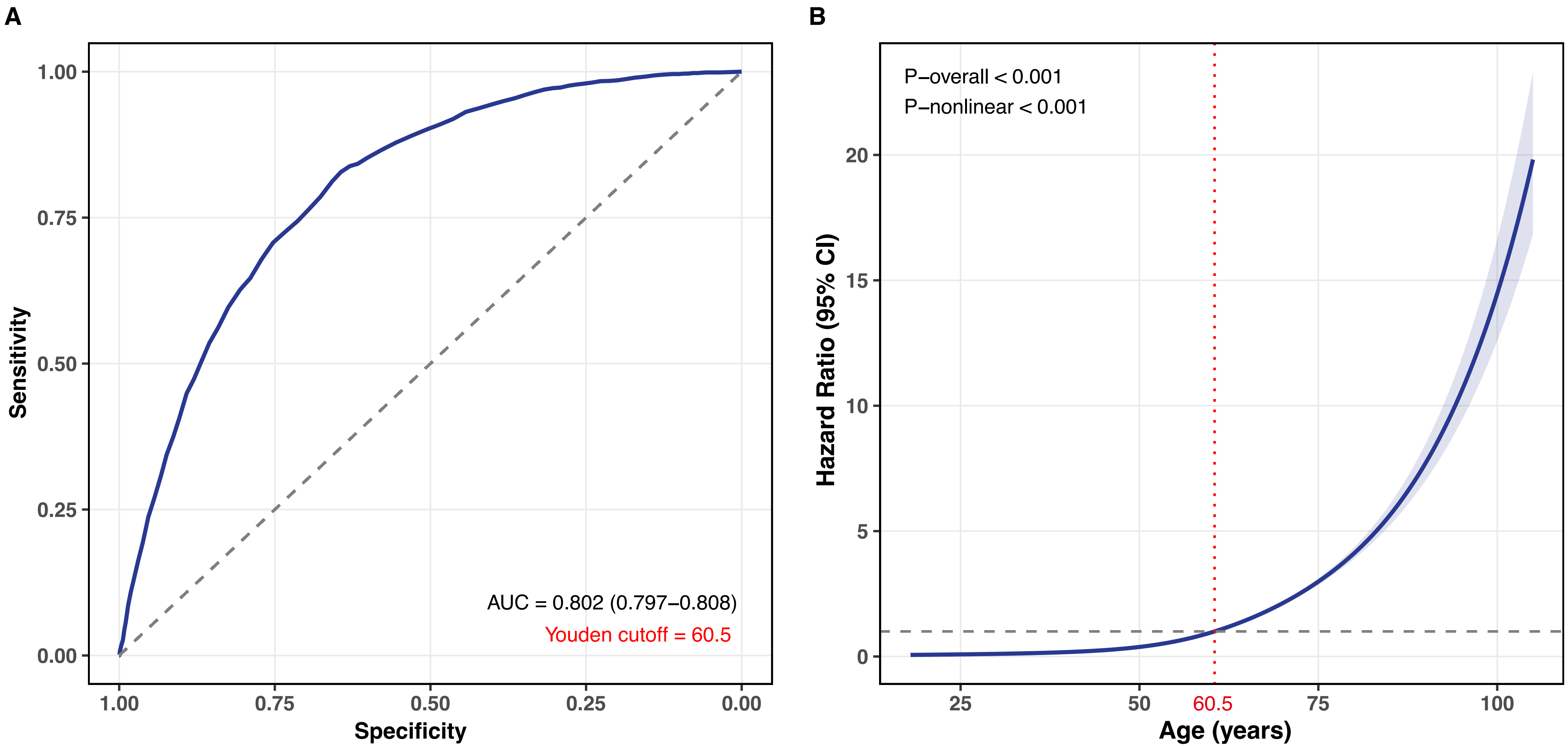

### Figure S2.pdf

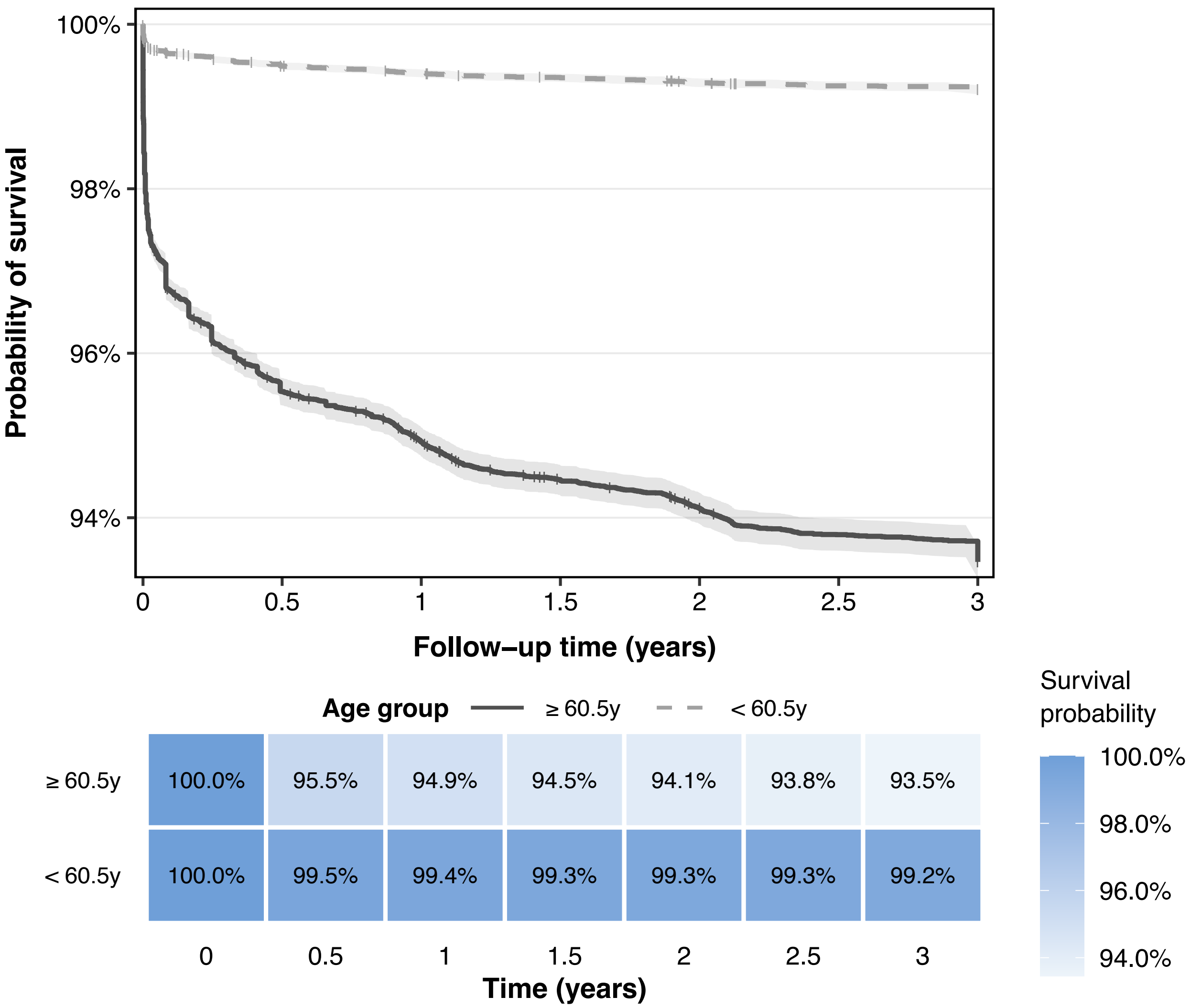

### Figure S3.pdf

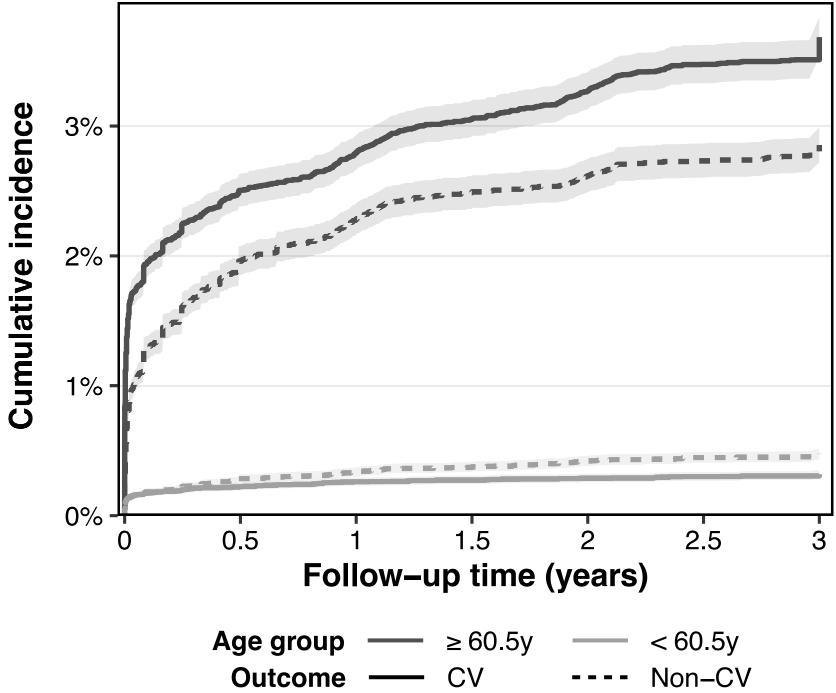


| | 0 | 0.5 | 1 | 1.5 | 2 | 2.5 | 3 |
|---|---|---|---|---|---|---|---|
| ≥ 60.5y I CV | 0.00% | 2.51% | 2.80% | 3.06% | 3.27% | 3.47% | 3.68% |
| ≥ 60.5y I Non–CV | 0.00% | 1.96% | 2.28% | 2.49% | 2.62% | 2.73% | 2.85% |
| < 60.5y I CV | 0.00% | 0.22% | 0.26% | 0.27% | 0.29% | 0.30% | 0.32% |
| < 60.5y I Non–CV | 0.00% | 0.29% | 0.34% | 0.38% | 0.42% | 0.45% | 0.48% |

Time (years)

Cumulative incidence: 0.0%, 1.0%, 2.0%, 3.0%

### Figure S4.pdf

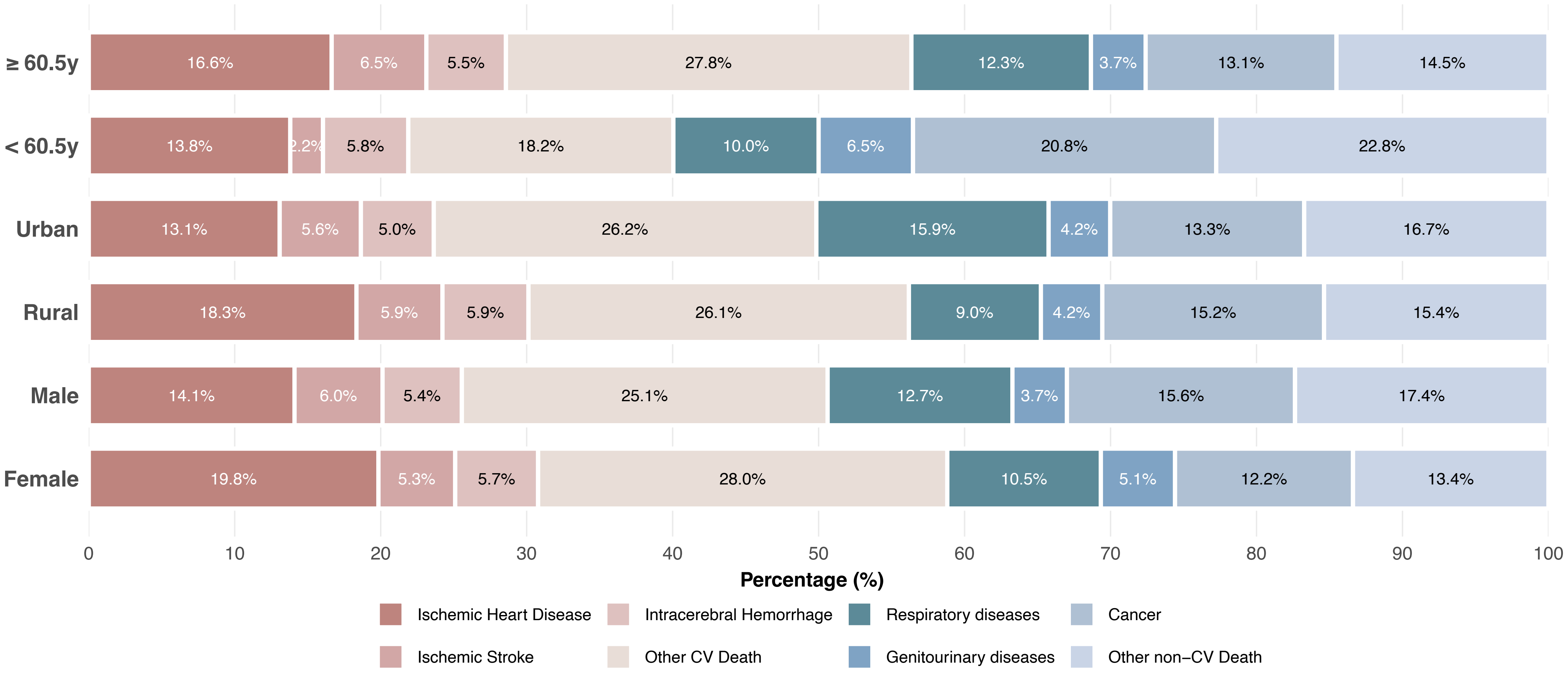
